# Identification and determination of the AST pattern of Acinetobacter species isolated from different clinical samples by VITEK □ Compact

**DOI:** 10.64898/2026.03.07.26347849

**Authors:** Pooja Rani, Nikhil Payal, Manisha Khandait, Swati Dixit

**Author notes:** **Author’s Information:**.

## Abstract

**Introduction:** *Acinetobacter* is a highly diverse genus which includes a range of common pathogenic species such as *A. baumannii, A. lwoffii* etc. *Acinetobacter* species causes bacteremia, pneumonia, wound infections, Urinary tract infections in community as well as hospital settings. *A. baumannii* is one of the ESKAPE pathogen which makes it even more lethal as antibiotics cannot action on this.

**Aim:** To isolate *Acinetobacter* species from various clinical samples and to check their antimicrobial susceptibility pattern by VITEK □ Compact in SGT Hospital, Gururam, Haryana.

**Results:** Out of total 6673 samples 595 were the positive isolates from which 35 were *Acinetobacter* isolates which were received from various wards of the hospital. Occurrence of *Acinetobacter* was seen more in males(57.14%) as compare to females (46.8%). A total of 31 strains were *A. baumannii*, 3 were *A. lwoffi* and 1 strain was of *A. haemolyticus*. Prominent presence of *Acinetobacter* was seen in Blood (48.57%) specimen along with pus(22.85%), endotracheal (22.85%), tracheal (2.85%) and eye swabs (2.85%). All the isolates were resistant to piperacillin/tazobactam (100%), ceftriazone (100%), amikacin (100%), gentamicin (100%) ciprofloxacin (91.42%), ceftazidime (91.42%), cefepime (88.57%), levofloxacin (88.57%) and trimethoprim/sulfamethoxazole (80%). Colistin susceptibility was observed in 88.57% of the isolates.

**Conclusion:** *Acinetobacter* is a common pathogen in hospital acquired as well as in community acquired infections as it is a opportunistic pathogen hence to identify the *Acinetobacter* species and to understand their antimicrobial resistance pattern this study was conducted.

## 1. INTRODUCTION

Genus *Acinetobacter* from Moraxellaceae family is a highly diverse aerobic, Gram negative coccobacilli which is saprophytic, oxidase positive and can form biofilm. Although *Acinetobacter* genus is highly diverse, there are more than 50 species known till date. However most of the species are widely dispersed in natural and clinical environment, and are non-pathogenic but species *A. baumannii, A. calcoaceticus* and *A. lwoffii* are most commonly known for causing infections. Species; *A. haemolyticus, A. johnsonii, A. junii, A. nosocomialis, A. pittii, A. schindleri*, and *A. ursingii* have also been reported as pathogens occasionally [1,2].

Most of the species of this genus are common pathogens causing community acquired infections as well as hospital acquired infections. Wound infections, urinary tract infections, pneumonia and bacteremia following trauma, urinary catheters, mechanical ventilators, and central venous access are among the infections associated with these species [3].

*Acinetobacter* species shoes resistance to wide variety of antibiotics because of low permeability of its outer cell membrane and it can also accumulate its resistance mechanisms encoded on integrons, plasmids and transposons [4,5].

*Acinetobacter baumannii* is one of the ESKAPE organisms (Enterococcus faecium, Staphylococcus aureus, *Klebsiella pneumoniae, A. baumannii, Pseudomonas aeruginosa, and Enterobacter spp*.), which was identified as priority 1 pathogen for new antimicrobial development by World Health Organisation (WHO) [4]. To emphasise that they escape the lethal action of antibiotics that pose a global threat to human health and a therapeutic challenge due to emerging and constantly increasing resistance [6,7]. As a health concern infections caused by *Acinetobacter* are difficult to treat because of the multidrug resistant strains and different antimicrobial resistance mechanism [4]. The aim of the study was to assess the identification and antimicrobial susceptibility pattern of this genus in various clinical samples by using VITEK □compact.

## 2. MATERIAL AND METHODS

The present study is reterospective cross sectional for over 1 year (1 Jan 2023 to 31 Dec 2023) and was conducted in the Department of Microbiology, Faculty of Medicine and Health Science, SGT University, Gurugram, India. A total of 6673 specimens were received in the Microbiology laboratory of SGT Hospital for culture and antimicrobial susceptibility from IPD and OPD which were included in this study. Blood, respiratory tract specimens, urine, pus, cerebrospinal fluid (CSF), tissue sample, plural fluid and other biological specimens collected were included in this study which were processed according to standard routine microbiology laboratory procedures. Urine samples were inoculated on CLED (cystein lysine electrolyte deficient) media, Blood specimens were processed in Bact/Alert for 5 days. Positive samples of blood further processed along with pus, eye swab, CSF, fluid and other clinical samples on Blood and MacConkey agar.

Identification of Gram negative bacteria was done by Gram’s staining, colony morphology on Blood agar, MacConkey agar and other biochemicals as Gram negative cocobacilli, non motile on motility test medium, oxidase negative and catalase postive, citrate positive and hemolysis on blood agar. *Acinetobacter* identification and antibiotic susceptibility was further confirmed by Vitek□compact. Piperacillin/Tazobactam, Ceftriazone, Cefoparazone/Sulbactum, Cefepime, Imepenem, Meropenem, Amikacin, Gentamicin, Ciprofloxacin, colistin and Trimethoprim/Sulfamethoxazole were the following antibiotics which were used to check the antimicrobial susceptibility of *acinetobacter* species, were used in VITEK card. From all 6673 clinical specimens 595 was culture positive isolates and out of 595 positive isolates 35 were reported as *acinatobacter*.

## 3. RESULTS

### 3.1 Isolation of Samples

A total of 6673 samples were received in microbiology laboratory of SGT Hospital from which only 595 were the positive isolates. Clinical samples were received from all the wards including IPD (49.24%), OPD (33.61%), ICU (5.8%), RICU (3.03%), SICU (2.86%), ICCU (2.02%), NICU (1.34%), HDU (0.84%), BICU (0.67%) and PICU (0.50%). The maximum number of positive isolates were obtained from IPD **(Table 1) (Fig. 1)**

**Figure 1.**
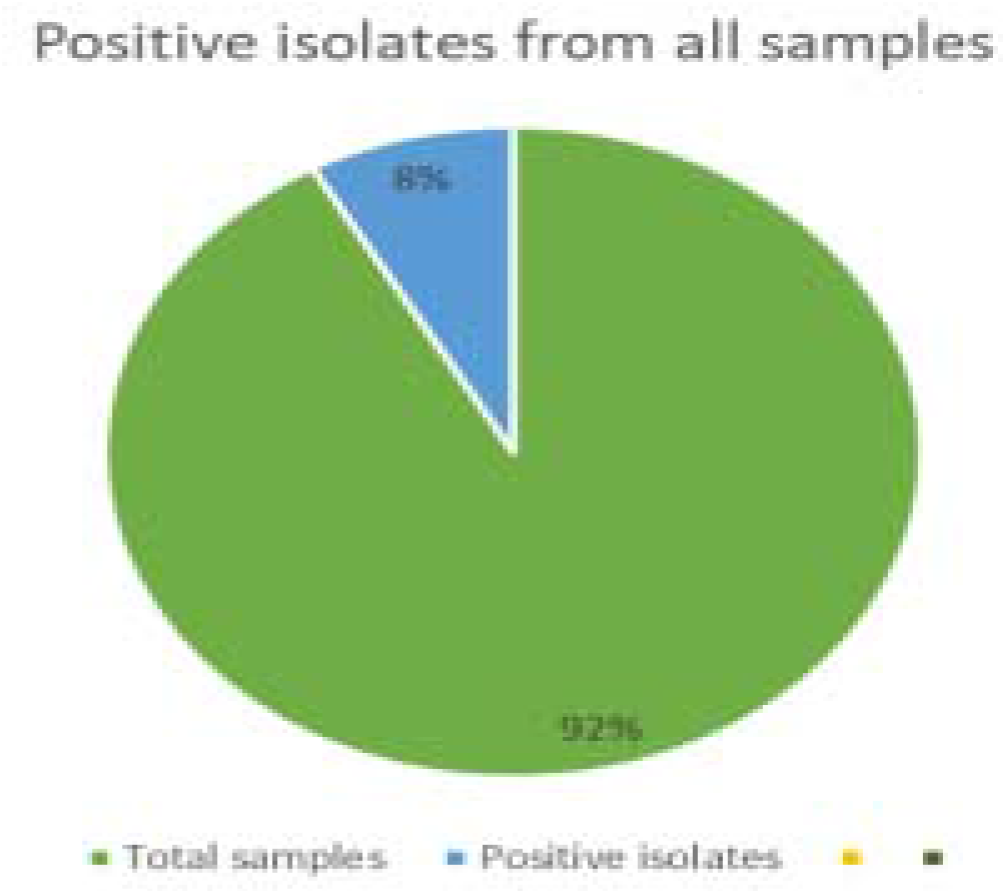
Chart representation showing the percentage of positive isolates from the total samples received from IPD

**Table 1.**
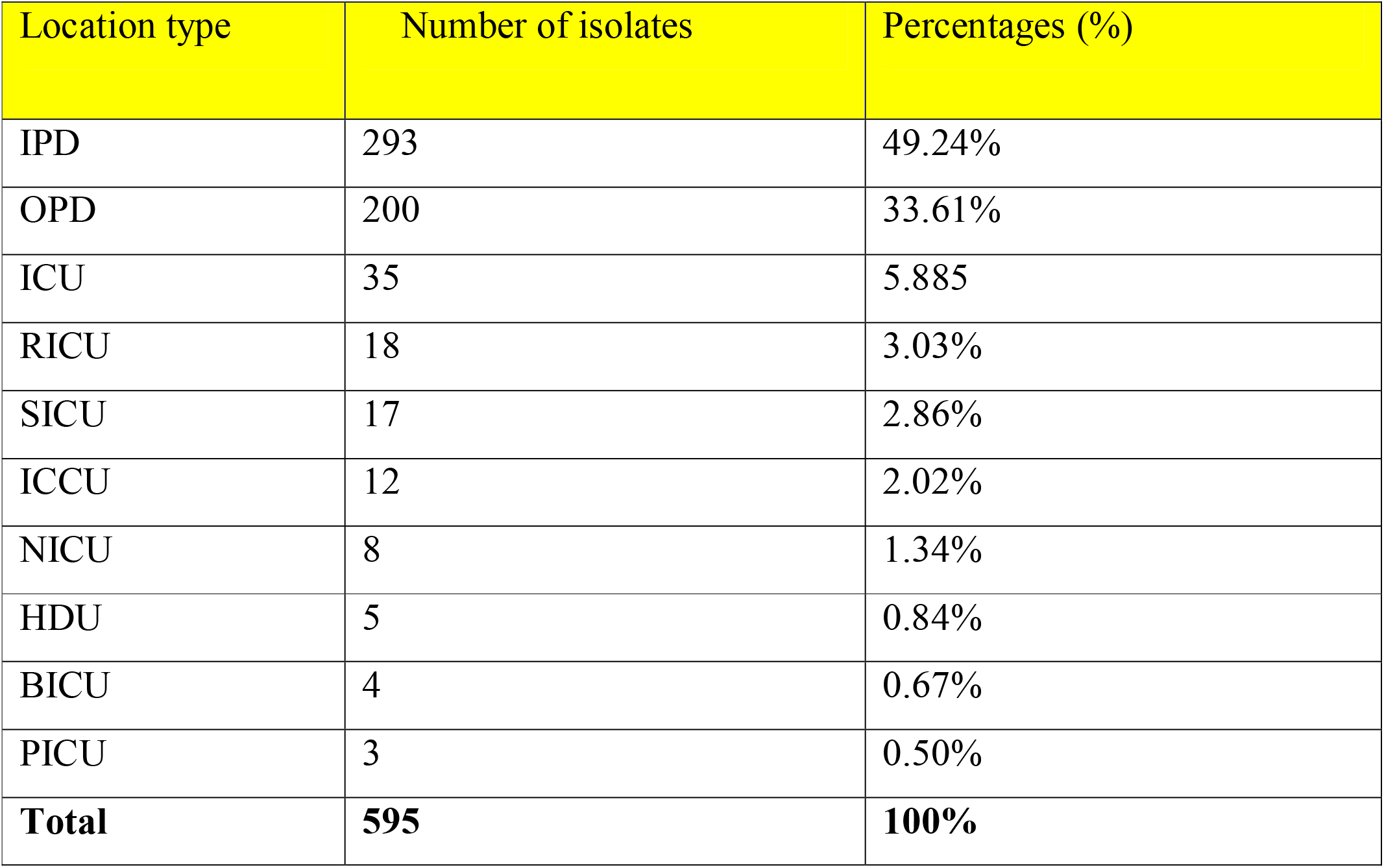
Number of isolates and their percentage of the clinical samples obtained from all the wards including IPD (49.24%), OPD (33.61%), ICU (5.8%), RICU (3.03%), SICU (2.86%), ICCU (2.02%), NICU (1.34%), HDU (0.84%), BICU (0.67%) and PICU (0.50%).

**Table 1a.**
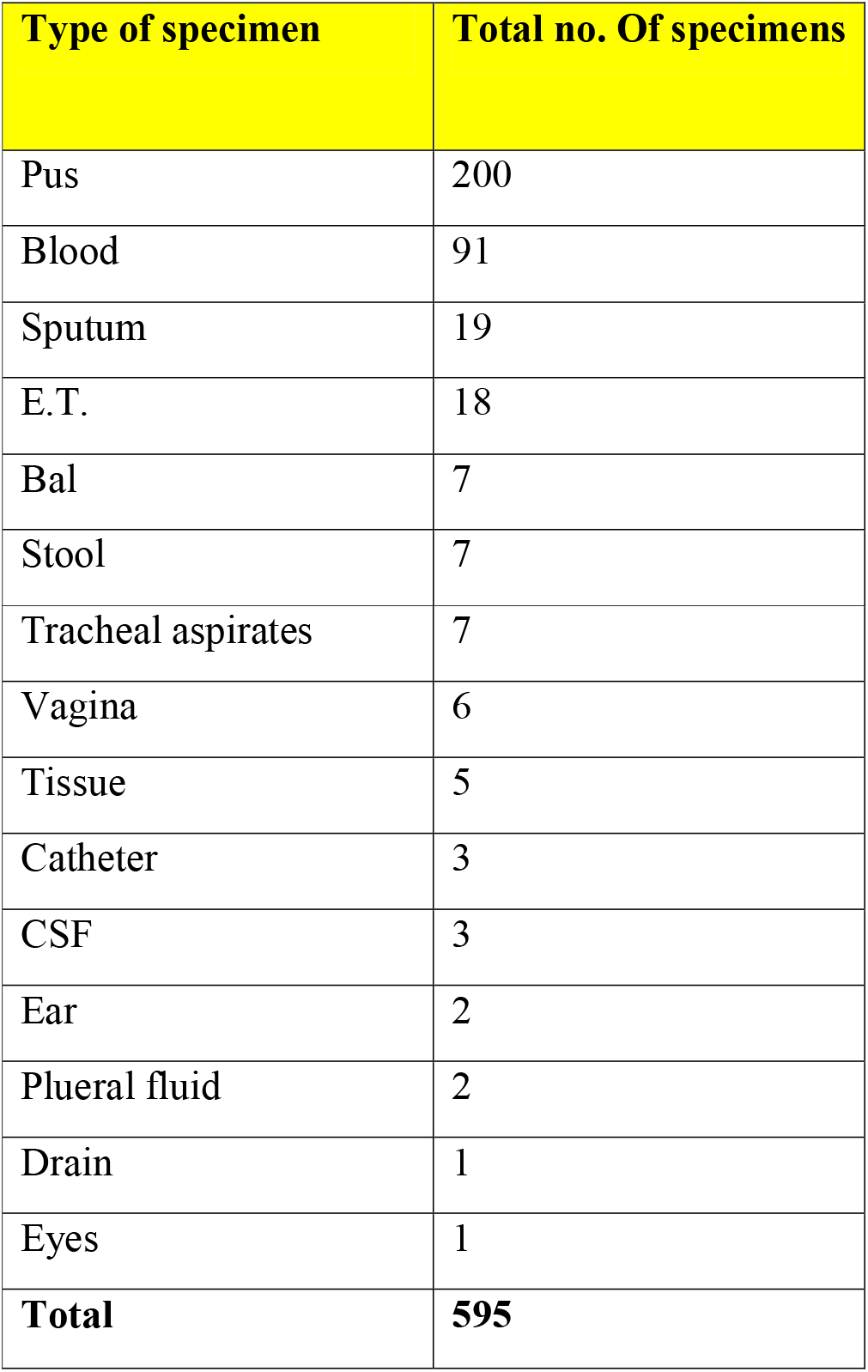
Number of isolates and their percentage of the clinical samples obtained from all the wards including including Urine (233), Pus (200), Blood (91), Sputum (19), Endotracheal (18), BAL (7), Stool (7), Tracheal aspirates (7), Vaginal swabs (6), Tissue (5), Catheter (3), CSF (3) Ear (2), Plueral fluid (2), Drain (1) and Eyes (1).

#### Observation

Out of 595 there were 35 isolates which were identified as *Acinetobacter* species, in different clinical samples. Pus was 7 (22.85%), Blood was 17 (48.57%), Endotracheal was 7 (22.85%), Tracheal aspirates was 1 (2.85%) and Eye swab was 1 (2.85%) were the prominent clinical samples for *acinetobacter* identification.

### 3.2 Positive isolates for Acinetobacter

**Table 3.**

| Speciemen type | No. Of positive isolates for acinetobacter | Percentage (%) |
| --- | --- | --- |
| Pus | 8 | 22.85% |
| Blood | 17 | 48.57% |
| E.T | 8 | 22.85% |
| Tracheal aspirate | 1 | 2.85% |
| Eye | 1 | 2.85% |

**Chart 2.**
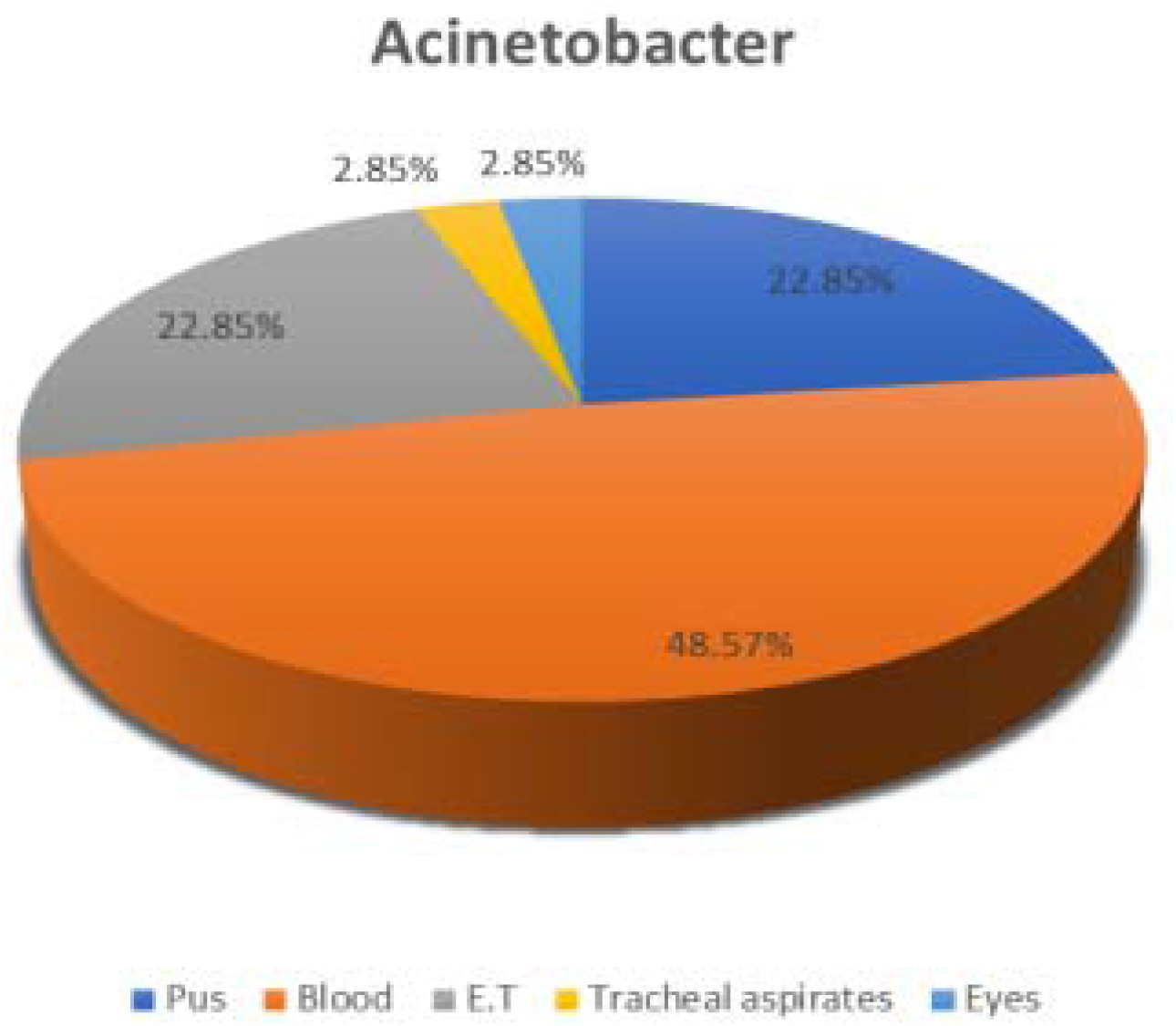

Out of all 35 isolates of *acinetobacter* 20 were males and 15 were females

**Chart 3.**
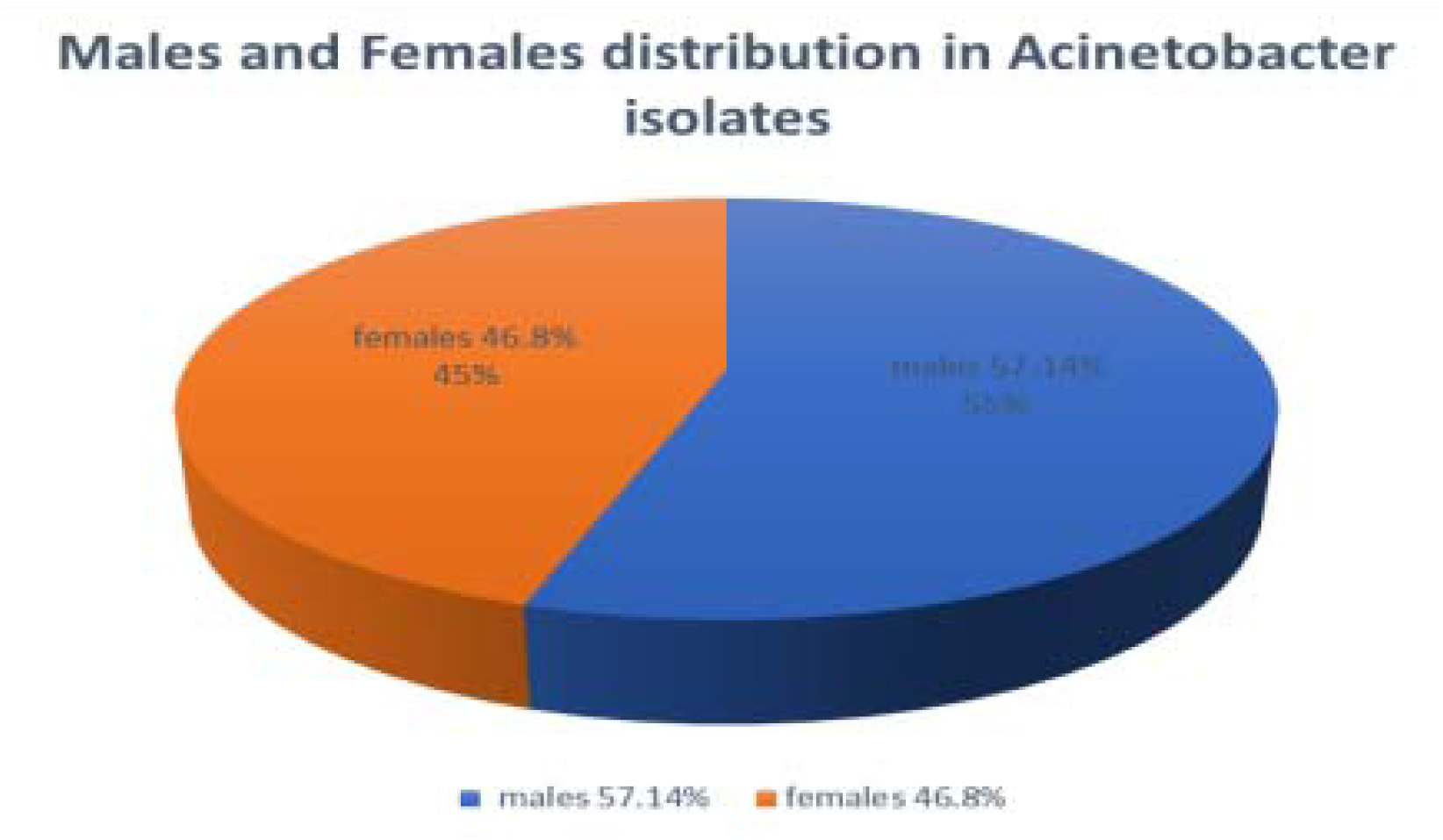

**Table 4.** Species distribution of *acinetobacter* isolates.

| *Acinetobacter* species | Number | Percentage |
| --- | --- | --- |
| *Acinetobacter baumannii* | 31 | 88.57% |
| *Acinetobacter lwoffii* | 3 | 8.57% |
| *Acinetobacter haemolyticus* | 1 | 2.85% |

#### Antibiotic susceptibility pattern

VITEK □Compact was used determination of antimicrobial resistivity and sensitivity pattern in *acinetobacter* species. Among all 35 isolates of *Acinetobacter* all shows 100% resistivity to pipeacil/tzobactam, ceftriazone, amikacin and gentamicin, High level of resistance were seen in ciprofloxacin (91.42%), ceftazidime (91.42%).0, cefepime (88.57%), levofloxacin (88.57%).

**Table 5.**

| Antibiotic | Sensitivity no. | Sensitivity % | Resistance No. | Resistivity % |
| --- | --- | --- | --- | --- |
| Piperacillin/tazobactam | 0 | 0% | 35 | 100% |
| Ceftriazone | 0 | 0% | 35 | 100% |
| Cefoperazone-sulbactam | 6 | 17.14% | 29 | 82.85% |
| Cefepime | 2 | 5.71% | 31 | 88.57% |
| Imepenam | 3 | 8.57% | 32 | 91.42% |
| Meropenam | 3 | 8.57% | 32 | 91.42% |
| Amikacin | 0 | 0% | 35 | 100% |
| Gentamicin | 0 | 0% | 35 | 100% |
| Ciprofloxacin | 3 | 8.57% | 32 | 91.42% |
| Colistin | 31 | 88.57% | 4 | 11.42% |
| Trimethoprim/sulfamethoxazole | 9 | 25.71% | 26 | 74.28% |
| ceftazidime | 3 | 8.57% | 32 | 91.42% |
| Levofloxacin | 4 | 11.42% | 31 | 88.57% |
| Minocycline | 13 | 37.14% | 5 | 14.28% |

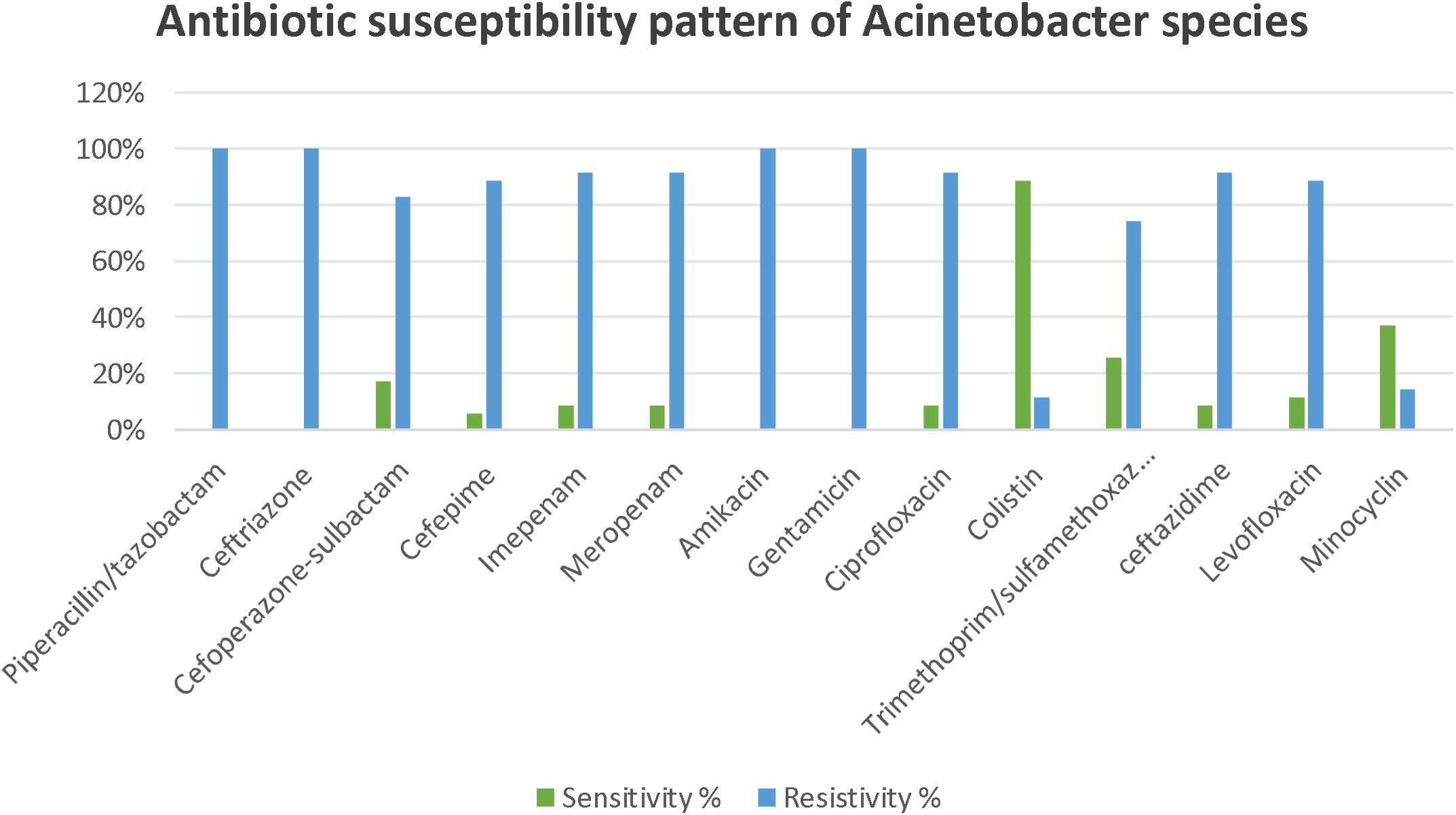

Among a total of 35 acinetobacter isolates 26 (74.28%) were MDR. All MDR isolates belonged to A. baumanii. Similar study was conducted in Nepal and Amatya et al reported 71.3% rate of MDR in Acinetobacter. Rajkumari S et al in her study reported 76.81% MDR Acinetobacter.

## Discussion

*Acinetobacter* is an nosocomial pathogen which is also an opportunistic pathogen commonly causing diseases in immnunocompromized patients and other patients. Genus *Acinetobacter* has become an important source of critical thinkimg due to its ability of showing resistance to most of the antibiotics present for treating the infections caused by this particular genus. *Acinetobacter* genus has emerged as a cause of ICU infections more overly.

We investigated the presence and antimicrobial susceptibility pattern of genus Acinetobacter and its species including *Acinetobacter baumannii, Acinetobacter lwoffii and Acinetobacter haemolyticus*; as these were the strains which were isolated from the different clinical samples which ere recived in the microbiology laboratory of SGT University, budhera, gurugram.

In our study we studied 35 isolates of *Acinetobacter* in which 31 were *Acinetobacter baumannii*, 3 were *Acinetobacter lwoffii* and 1 was *A. haemolyticus*.

In our study the prevalence of *Acinetobacter* was 5.88% which was comparable with a study done by RB Madhavi et al which was 8.9%.^8^ and Chitrabhanu et al who reported as 6.2%.^9^ Majority of *Acinetobacter* infections in our study was isolated from blood specimens (48.57%) and in a similar study SH Mohammed shows 36.9% prevalence of *Acinetobacter* in blood infections. ^**10**^

In our study, Infections due to *Acinetobacter* were seen more common in males(57.14%) as comparative to females (46.8%), similar findings were also observed by RB Madhavi *Acinetobacter* infections occurred significantly in male patients (65.7%) than in female patients (34.3%). ^8^

In our study specimens which commonly yielding growth for *acinetobacteria* was blood (48.57%), pus (22.85%), Endotracheal (22.85%), tracheal aspirates (2.85%) and eye swab (2.85%).

In our study *Acinetobacter baumannii* (80%) is prominently present in most of the clinical specimens along with *A. lwoffii* shows 14.28% and *A. haemolyticus* shows 5.71% prevalence among all the isolates reported positive for *Acinetobacter*. Similar study was also conducted by R Kaur et al Acinetobacter spp. Reported in their study was, 91.6% were *Acinetobacter Baumanii &* 5.6% *Acinetobacter lwoffi*, 2.8% *Acinetobacter hemolyticus*.^11^ Therefore *A. baumannii* was prominent in all clinical specimens which were positive for *Acinetobacter*, this was possible due to three major factors which may contributing to the persistence a of *A. baumannii* i.e., because of high level of resistance to major antimicrobials, suitable environment and immunocompromised patients.^12^

High level of resistance were seen for piperacillin/tazobactam (100%), ceftriazone (100%), amikacin (100%), gentamicin (100%) ciprofloxacin (91.42%), ceftazidime (91.42%), cefepime (88.57%), levofloxacin (88.57%) and trimethoprim/sulfamethoxazole (80%) in our study. Taneja et al in their study reported resistance of *Acinetobacter* towards Gentamicin and ciprofloxacin was 79.5% and 72.8% respectively.^13^ Resistivity towards imepenam (62.85%), meropenem (54.28%) and minocycline (14.28%) were reported. Amardeep et al in a similar study reported the resistance of *Acinetobacter*, towards imipenem (42.6%) and meropenem (55.4%).^14^ High Sensitivity towards colistin (42.85%) were reported among all the isolates of *Acinetobacter* genus. Similar study was conducted by P Singh et al showing 92.6 % sensitivity of *Acinetobacter* isolates towards colistin.^15^

## Conclusion

Our study showed the significant prevalence of *acinetobacter* in our area (SGT hospital, gurugram; Delhi NCR region). 5.88% prevalence along with 80.4% of resistance towards antibiotics were reported in our study which is a challenge for healthcare system to overcome because of high resistance towards commonly used antibiotics, however colistin is somehow showing an emerging sensitive effect on *acinetobacter*.Regular use of antibiotics without proper investigations is leading to resistance of commonly used antibiotics for treating infections. This indicates the role of *acinetobacter* in nosocombial and opportunistic pathogen causing a wide variety of infections.

Aim of our study was to isolate *acinetobacter* species and to report the antimicrobial resistance pattern of genus *acinetobacter* from various clinical samples in our hospital. Hence this study was conducted to strengthening hospital infection control and prevention control practices in health care settings and projecting attention to the use of antimicrobials accordingly which may helps in preventing the occurrence of MDR *acinetobacter* species.

## Data Availability

All data produced in the present work are contained in the manuscript

